# Validation of a new automated chemiluminescent anti-SARS-CoV-2 IgM and IgG antibody assay system detecting both N and S proteins in Japan

**DOI:** 10.1101/2020.07.16.20155796

**Authors:** Rin Yokoyama, Makoto Kurano, Yoshifumi Morita, Takuya Shimura, Yuki Nakano, Chungen Qian, Fuzhen Xia, Fan He, Yoshiro Kishi, Jun Okada, Naoyuki Yoshikawa, Yutaka Nagura, Hitoshi Okazaki, Kyoji Moriya, Yasuyuki Seto, Tatsuhiko Kodama, Yutaka Yatomi

**Affiliations:** Department of Clinical Laboratory, the University of Tokyo Hospital, Tokyo, Japan; Department of Clinical Laboratory Medicine, Graduate School of Medicine, the University of Tokyo, Tokyo, Japan; The Key Laboratory for Biomedical Photonics of MOE at Wuhan National Laboratory for Optoelectronics - Hubei Bioinformatics & Molecular Imaging Key Laboratory, Systems Biology Theme, Department of Biomedical Engineering, College of Life Science and Technology, Huazhong University of Science and Technology, Hubei, P.R. China; Reagent R&D Center, Shenzhen YHLO Biotech Co., Ltd, Guangdong, P.R. China; Business Planning Department, Sales & Marketing Division, Medical & Biological Laboratories Co., Ltd, Tokyo, Japan; Department of Blood Transfusion, the University of Tokyo Hospital, Tokyo, Japan; Department of Infection Control and Prevention, The University of Tokyo, Tokyo, Japan; Department of Gastrointestinal Surgery, The University of Tokyo, Japan; Laboratory for Systems Biology and Medicine, The University of Tokyo, Tokyo, Japan

**Author notes:** **Address correspondence to:** Makoto Kurano, MD, PhD and Yutaka Yatomi, MD, PhD, Department of Clinical Laboratory Medicine, Graduate School of Medicine, The University of Tokyo, 7-3-1 Hongo, Bunkyo-ku, Tokyo 113-8655, Japan, (M.K.), (Y.Y).

## Abstract

PCR methods are presently the standard for the diagnosis of Coronavirus disease 2019 (COVID-19), but additional methodologies are needed to complement PCR methods, which have some limitations. Here, we validated and investigated the usefulness of measuring serum antibodies against severe acute respiratory syndrome coronavirus 2 (SARS-CoV-2) using the iFlash3000 CLIA analyzer. We measured IgM and IgG titers against SARS-CoV-2 in sera collected from 26 PCR-positive COVID-19 patients, 53 COVID-19-suspected but PCR-negative patients, and 20 and 100 randomly selected non-COVID-19 patients who visited our hospital in 2020 and 2017, respectively. The within-day and between-day precisions were regarded as good, since the coefficient variations were below 5%. Linearity was also considered good between 0.6 AU/mL and 112.7 AU/mL for SARS-CoV-2 IgM and between 3.2 AU/mL and 55.3 AU/mL for SARS-CoV-2 IgG, while the linearity curves plateaued above the upper measurement range. We also confirmed that the seroconversion and no-antibody titers were over the cutoff values in all 100 serum samples collected in 2017. These results indicate that this measurement system successfully detects SARS-CoV-2 IgM/IgG. We observed four false-positive cases in the IgM assay and no false-positive cases in the IgG assay when 111 serum samples known to contain autoantibodies were evaluated. The concordance rates of the antibody test with the PCR test were 98.1% for SARS-CoV-2 IgM and 100% for IgG among PCR-negative cases and 30.8% for SARS-CoV-2 IgM and 73.1% for SARS-CoV-2 IgG among PCR-positive cases. In conclusion, the performance of this measurement system is sufficient for use in laboratory testing.

## Introduction

In December 2019, the first pneumonia cases caused by an unknown microorganism were identified in Wuhan, China.[1] The pathogen was identified as a novel betacoronavirus and was named “severe acute respiratory syndrome coronavirus 2 (SARS-CoV-2). “[2] SARS-CoV-2 is phylogenetically similar to SARS-CoV, which caused outbreaks of a severe respiratory syndrome in China in 2002.[3] The symptoms of coronavirus disease 2019 (COVID-19), which is the respiratory syndrome caused by SARS-CoV-2, are fever, cough and lymphopenia.[4] Chest computed tomography examinations of COVID-19 patients are characterized by the bilateral distribution of patchy shadows or ground-glass opacities.[5] Since early December 2019 and as of June 15, 2020, over 7,800,000 cases of COVID-19 have been confirmed and 430,000 deaths have been reported throughout the world,[6] and the World Health Organization has reported a fatality rate for cases defined as pneumonia of approximately 2%.[7]

Currently, COVID-19 is diagnosed by the clinical presentation of the patient, as described above, and the detection of SARS-CoV-2 RNA in respiratory specimens, such as a nasal swab or sputum, using real-time reverse-transcription polymerase chain reaction (RT-PCR).[8,9] However, this method requires skilled technicians who know how to handle genetic samples and perform PCR tests and occasionally causes false-negative results because of the viral replication window, a low viral titer, or incorrect sample collection.[10] Moreover, the sampling of respiratory specimens exposes medical staff to a higher risk of secondary infection through aerosolization than the sampling of sera.[11,12] Therefore, other methods are required to complement PCR testing.

Candidate complementary tests include both antigen and antibody tests. Regarding antigen tests, although this method does not require special skills for performing genetic testing, there remains a high risk of secondary infection during sampling, and the sensitivity of antigen tests is reportedly lower than that of PCR testing.[13,14] Antibody tests are another candidate. Compared with PCR testing, this serological test method has a faster turn-around time and requires easier and safer sample collection and less specialized skills for technicians; furthermore, when we interpret the results of an antibody testing considering the duration after the onset of COVID-19, this test would give us important information in diagnosing COVID-19. The main concern regarding antibody tests is the high frequency of false-positive cases, which is a parameter that should depend on the quality of the test product.[15,16] Recently, Shenzhen YHLO Biotech Co., Ltd (China) has developed an antibody test with a high specificity[17–19]; however, this method has not yet been validated in the Japanese population. Therefore, in the present study, we aimed to validate the measurement of IgM and IgG antibodies against SARS-CoV-2 in sera and to investigate the usefulness of this method for the diagnosis of COVID-19.

## Materials and Methods

### Subjects

We enrolled a total of 26 COVID-19-positive cases and 53 COVID-29-suspected cases who were hospitalized at The University of Tokyo Hospital. Confirmed COVID-19 cases were defined based on the guidelines of The University of Tokyo Hospital. Briefly, patients with acute respiratory infection syndrome accompanied by detectable SARS-CoV-2 RNA in a throat swab or sputum at least once were confirmed as having COVID-19 (PCR-positive cases). Suspected patients were defined as subjects with respiratory symptoms, a history of overseas travel, or a high-risk contact with a confirmed COVID-19 case but with negative PCR results. We collected residual sera available after routine clinical testing and kept it frozen until measurement. The serological tests were performed using the sample that had been collected on the day closest to the day on which the sample for the PCR test had been collected. As control groups, we randomly selected 20 and 100 outpatients who had visited The University of Tokyo Hospital in March 2020 or January-December 2017, respectively. We also corrected 111 serum samples known to contain autoantibodies.

The current study was performed in accordance with the Declaration of Helsinki. Informed consent was obtained in the form of an opt-out form on the institution’s website. This study was conducted with the approval of The University of Tokyo Medical Research Center Ethics Committee (2019300NI-3).

### Antibody testing

Antibody testing was performed using SARS-CoV-2 IgM and IgG chemiluminescence immunoassay (CLIA) kits supplied by Shenzhen YHLO Biotech Co., Ltd. (China) and an iFlash3000 fully automated CLIA analyzer also from Shenzhen YHLO Biotech Co., Ltd. (China). Two antigens of SARS-CoV-2 (nucleocapsid protein [N protein] and spike protein [S protein]) were coated on the magnetic beads of these CLIA assays. The SARS-CoV-2 IgM or IgG titers were calculated as relative light units (RLU) obtained from the analyzer and were described as arbitrary units per milliliter (AU/mL). According to the manufacturer’s instructions, the cutoff value for a positive SARS-CoV-2 IgM/IgG result was deemed as 10 AU/mL. If the SARS-CoV-2 IgG titer was over 40 AU/mL, the sample was diluted with non-reactive serum and the antibody titers were measured once again.

### Method validation

The within-day precision was validated by measuring 3 kinds of serum samples with 20 replications simultaneously. Then, we calculated the mean value, the standard deviation (SD), and the coefficient of variation (CV) for each sample. The between-day precision was validated using two kinds of quality-control samples provided by the manufacturer. These samples were measured twice a day on days 0, 1, 2, 3, 4, 5, 9-11, and 16. Linearity was investigated using two kinds of pooled serum samples. Briefly, each sample was diluted with pooled non-reactive serum in two-fold serial dilutions for ten times. To investigate the existence of the prozone phenomenon, we diluted the samples with high concentrations of SARS-CoV-2 IgM and IgG titers using ten-fold serial dilutions for the SARS-CoV-2 IgM assay and two-fold serial dilutions for the SARS-CoV-2 IgG assay.

### Statistical analysis

The data were analyzed using StatFlex software (Osaka, Japan). The results were expressed as the mean ± SD. The Dunn test was used for comparisons of antibody titers between the control and other groups. A value of *p* < 0.05 was regarded as statistically significant in all the analyses.

## Results

### Precision and accuracy of antibody testing

First, we investigated the precision and accuracy of the antibody testing. As shown in Table 1A, the intra-day precision values were 1.9%-2.9% for SARS-CoV-2 IgM and 1.1%-3.3% for SARS-CoV-2 IgG, while the between-day precision values were 1.9%-3.3% for SARS-CoV-2 IgM and 2.5%-3.7% for SARS-CoV-2 IgG.

**Table 1.** Within-day and between-day precision of SARS-CoV-2 IgM/IgG titers.

| **A. Within-day precision** |  |  |  |  |
| --- | --- | --- | --- | --- |
|  | Sample | Mean<br>(AU/mL) | SD (AU/mL) | CV (%) |
| SARS-CoV-2 IgM | 1 | 2.0 | 0.04 | 2.03 |
|  | 2 | 7.0 | 0.13 | 1.86 |
|  | 3 | 83.8 | 2.46 | 2.94 |
| SARS-CoV-2 IgG | 4 | 3.7 | 0.06 | 1.68 |
|  | 5 | 10.2 | 0.11 | 1.06 |
|  | 6 | 74.3 | 2.47 | 3.32 |
| **B. Between-day precision** |  |  |  |  |
|  | Sample | Mean<br>(AU/mL) | SD (AU/mL) | CV (%) |
| SARS-CoV-2 IgM | 1 | 3.8 | 0.13 | 3.31 |
|  | 2 | 15.9 | 0.30 | 1.90 |
| SARS-CoV-2 IgG | 3 | 5.1 | 0.13 | 2.53 |
|  | 4 | 17.5 | 0.64 | 3.67 |

(A) Within-day precision. We measured SARS-CoV-2 IgM and IgG in three kinds of samples; each sample was analyzed with 20 replicates on the same day. (B) Between-day precision. We continuously measured two kinds of samples on days 0, 1, 2, 3, 4, 5, 9-11, and 16; each sample was analyzed with 2 replicates.

### Measurement range of SARS-CoV-2 antibody testing

To explore the measurement range of this antibody test, we performed a linear regression analysis. When we investigated linearity using samples with moderate antibody titers, the curves showed a good linearity between 0.6 AU/mL and 112.7 AU/mL for SARS-CoV-2 IgM and between 3.2 AU/mL and 55.3 AU/mL for SARS-CoV-2 IgG (Fig. 1). Next, we measured samples with high antibody titers to determine the upper limit of the measurement range. In the SARS-CoV-2 IgM assay, the upper curve increased up to a value of 2,405 AU/mL and then reached a plateau at higher concentrations. In the SARS-CoV-2 IgG assay, the curve reached a plateau at values over 73 AU/mL (Fig. 2). Therefore, we diluted the samples and determined the SARS-CoV-2 IgG titers for SARS-CoV-2 IgG levels over 50 AU/mL. In both assays, a hook effect was not observed.

**Fig 1.**
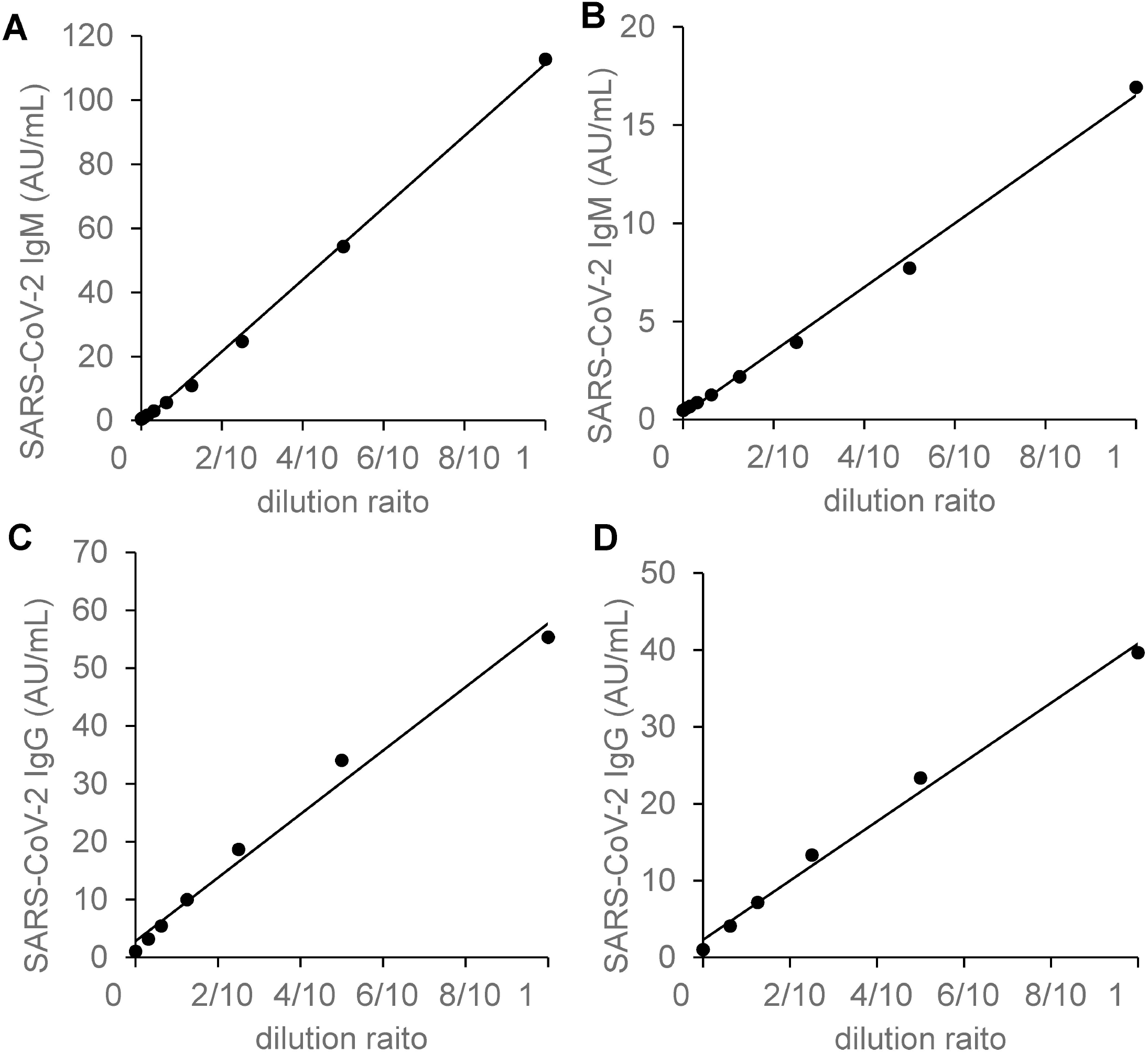
Linearity analyses of SARS-CoV-2 antibody titer. The dilution linearities of SARS-CoV-2 IgM (A, B) and SARS-CoV-2 IgG (C, D) were investigated. A sample was diluted with non-reactive serum in 5 to 8 steps; each sample was then analyzed with two replicates.

**Fig 2.**
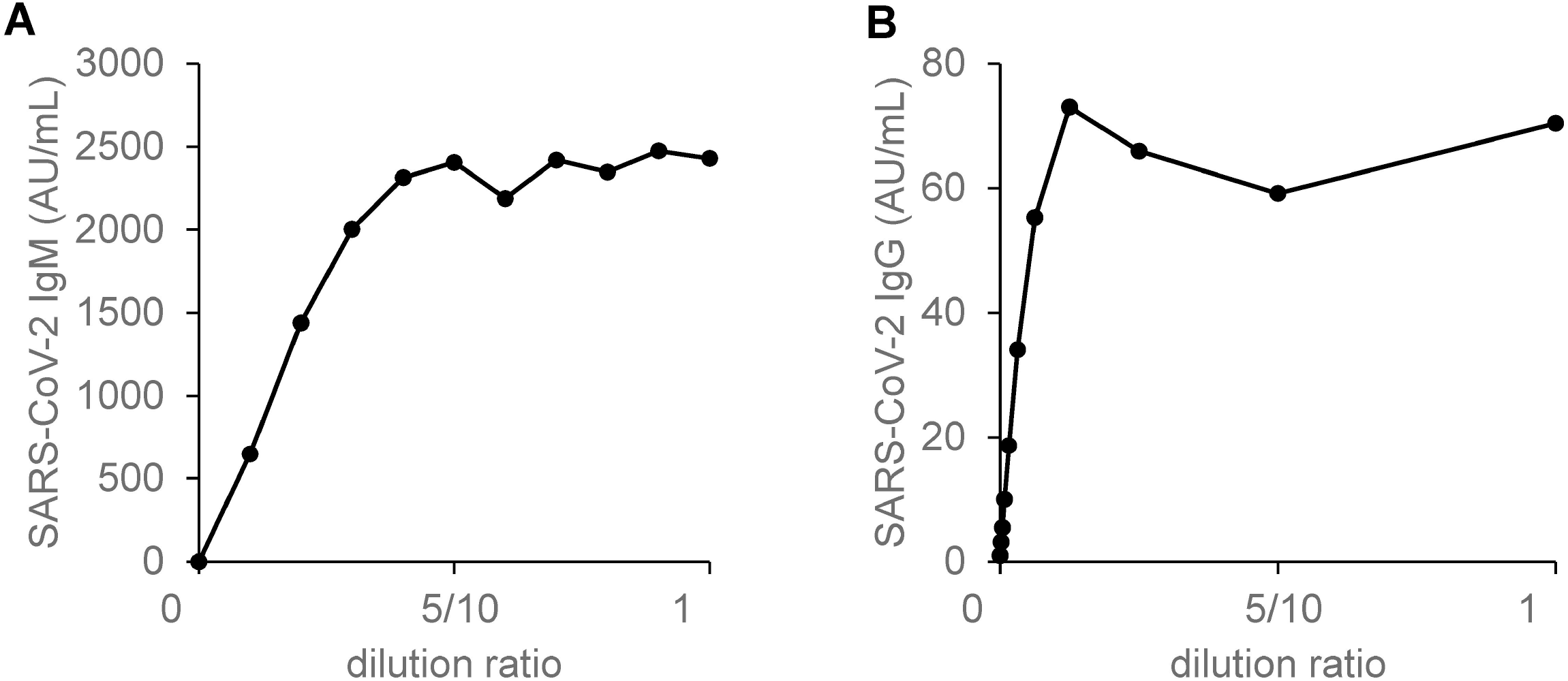
Prozone phenomena of SARS-CoV-2 antibody titer. The prozone phenomena of SARS-CoV-2 IgM (A) and SARS-CoV-2 IgG (B) were investigated. We diluted two serum samples from infected patients with non-reactive serum in 10 steps; each sample was then analyzed with two replicates.

### Successful detection of SARS-CoV-2 IgM/IgG

To confirm that this antibody measurement system could detect SARS-CoV-2 IgM/IgG successfully, we measured the antibody titers in sera obtained before and after infection with SARS-CoV-2 in three cases of COVID-19 confirmed using RT-PCR tests. As shown in Fig. 3, SARS-CoV-2 IgM and IgG were not detected before symptom onset; at several days after symptom onset, tests for both antibodies became positive and the titers gradually increased. In case 1, the IgM test once again became negative 19 days after hospitalization.

**Fig 3.**
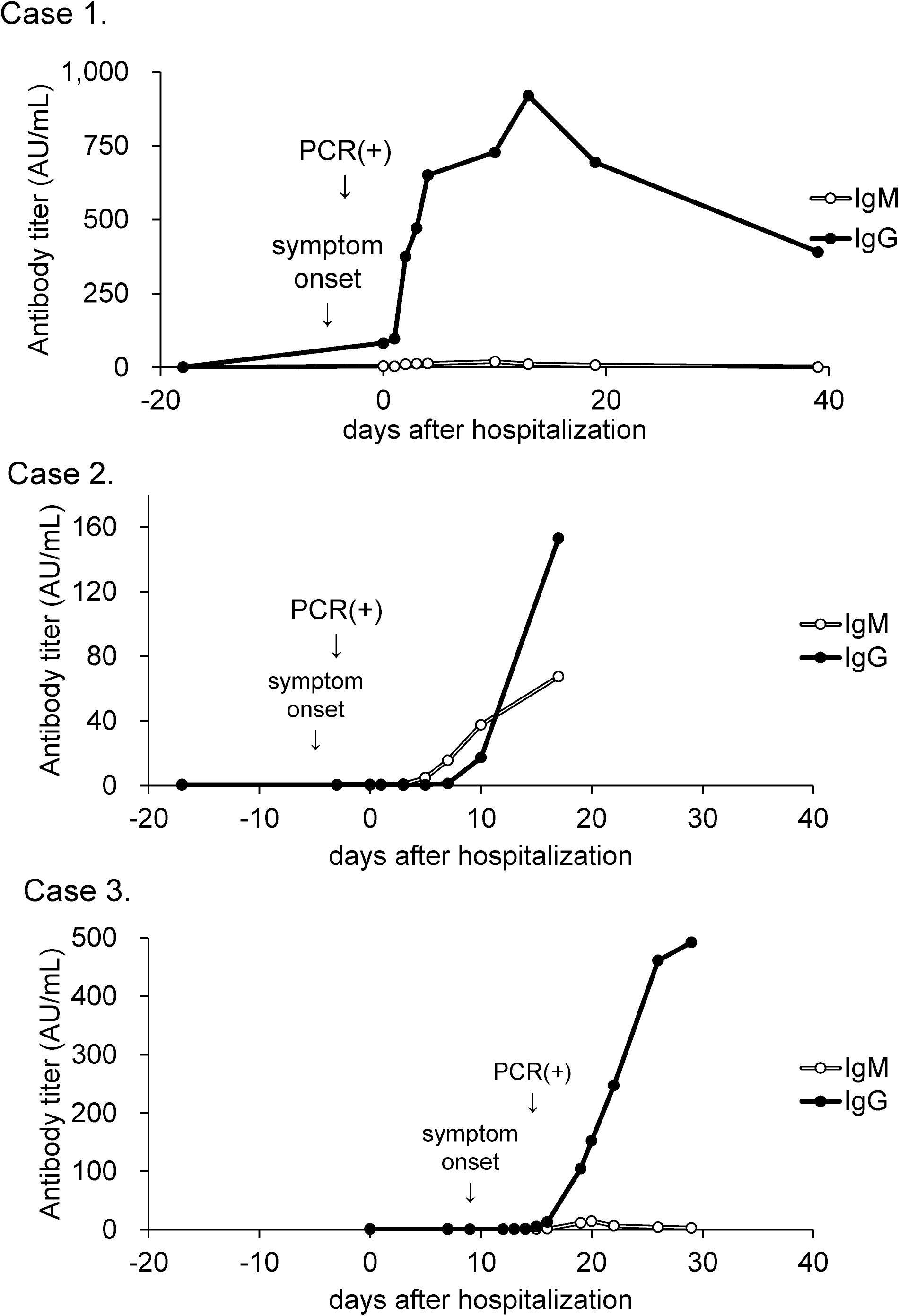
Time course of serum antibody titers in COVID-19 subjects. The time courses of the SARS-CoV-2 IgM and SARS-CoV-2 IgG titers in sera collected before and after the onset of COVID-19 were examined in three patients.

Second, we obtained 100 random serum samples collected from outpatients who had visited The University of Tokyo Hospital in 2017, when SARS-CoV-2 did not exist. None of these samples had an antibody titer over 10 AU/mL, suggesting that this measurement system can detect SARS-CoV-2 IgM/IgG without false-positive results (Fig. 4).

**Fig 4.**
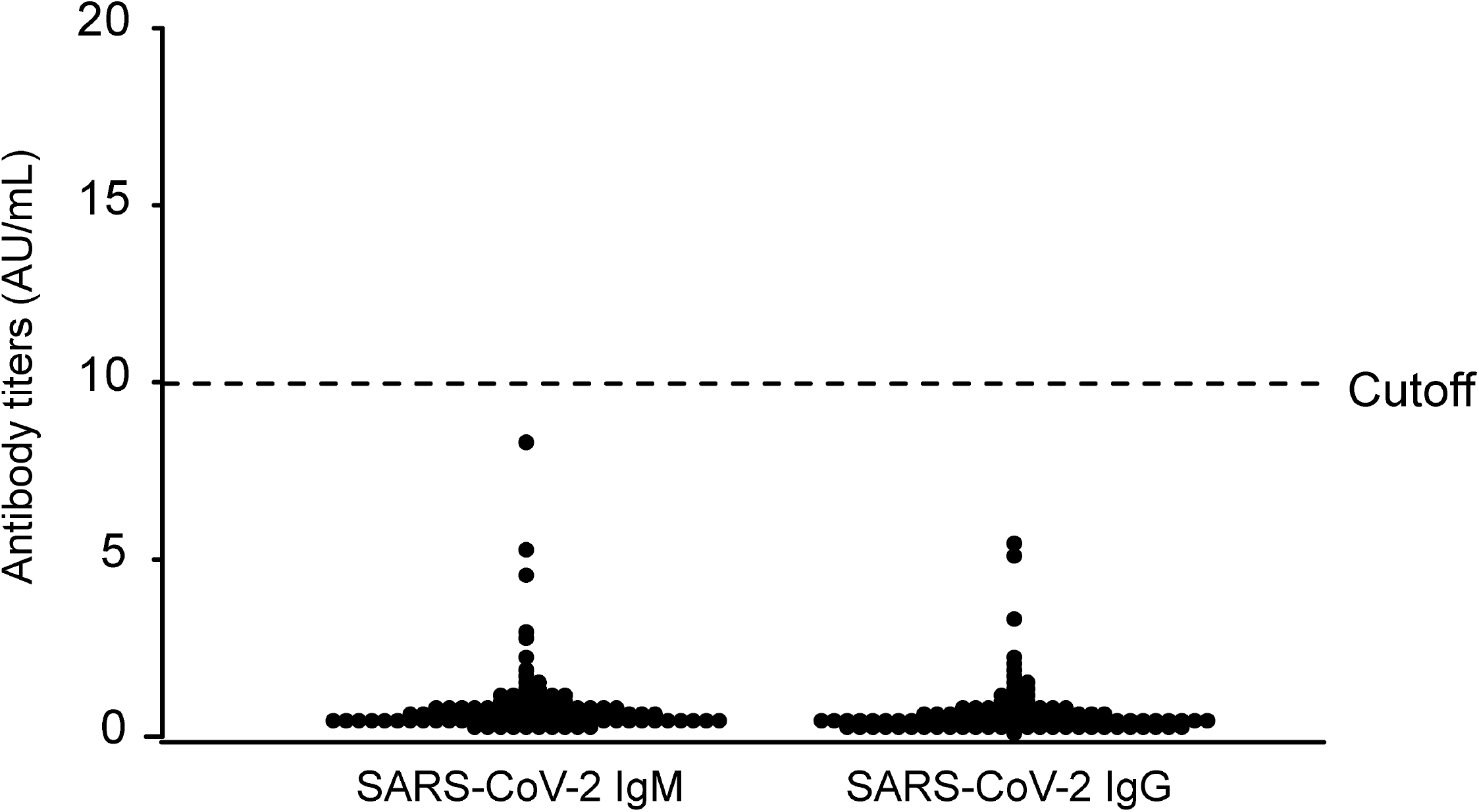
Serum antibody titers in sera from 2017. The SARS-CoV-2 IgM/IgG titers of sera collected from subjects (n = 100) in 2017 were measured.

### Cross-reactivity with autoantibodies

Since the presence of autoantibodies can sometimes affect the results of serological tests, we measured SARS-CoV-2 IgM/IgG in residual serum samples collected from patients with one of 5 different autoimmune diseases. For IgM, most of the serum samples from the patients with autoimmune diseases did not have a result over 10 AU/mL. However, two rheumatoid factor-positive patients, one anti-double-strand DNA antibody-positive patient, and one anti-mitochondrial M2 antibody-positive patient had values that exceeded the cutoff value (Fig. 5A). For IgG, none of the autoantibody-positive serum samples had a result that was over 10 AU/mL (Fig. 5B).

**Fig 5.**
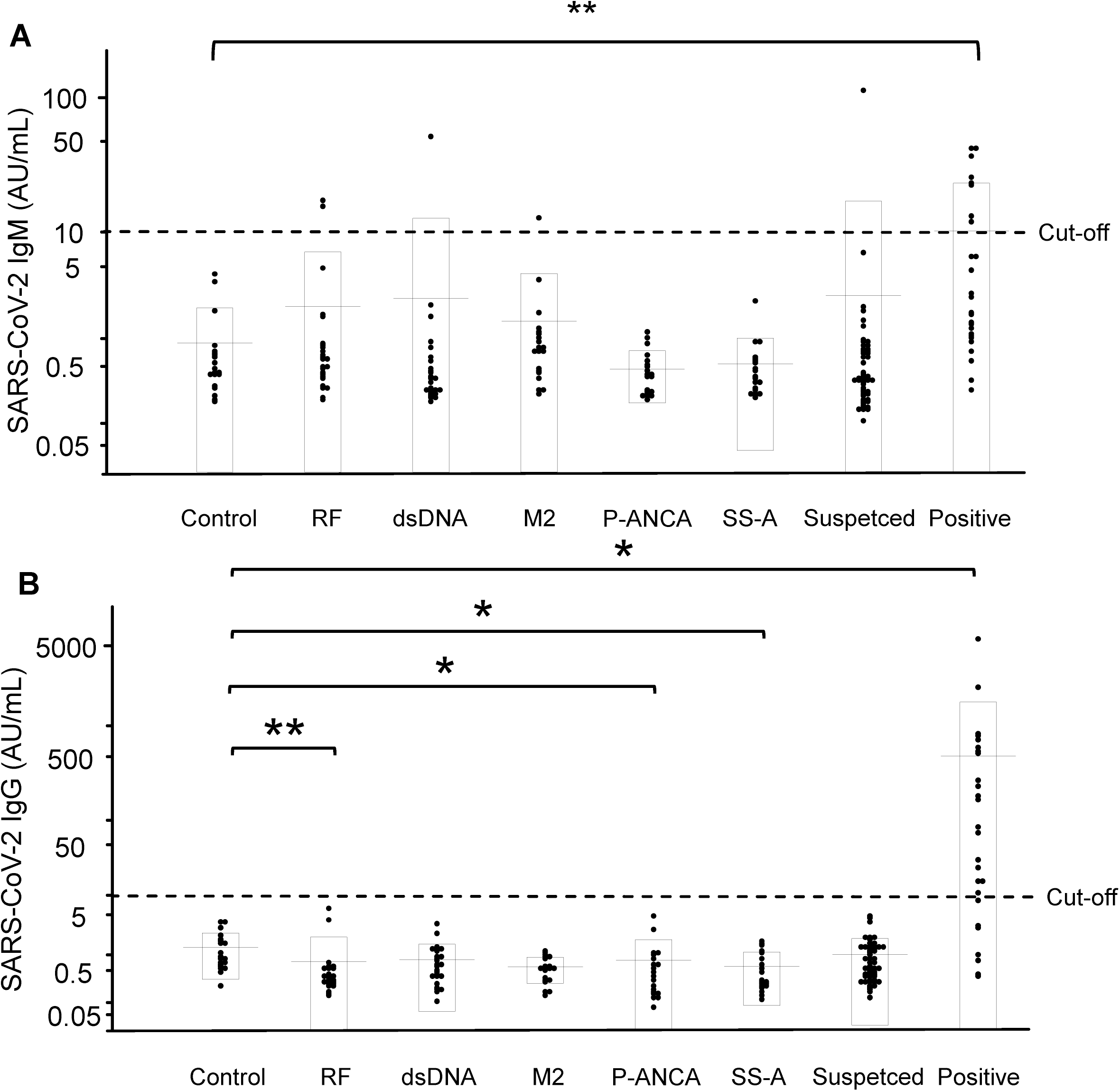
Interference from autoantibodies in SARS-CoV-2 IgM/IgG assay. We collected sera from patients with autoimmune diseases and measured the SARS-CoV-2 IgM (A) and IgG (B) titers. \**p* < 0.05, \*\**p* < 0.01. Control, randomly selected outpatients who visited the hospital in 2020 (n = 20); RF, rheumatoid factor-positive group (n = 25); dsDNA, anti-double-strand DNA antibody-positive group (n = 26); M2, anti-mitochondrial M2 antibody-positive group (n = 20); P-ANCA, myeloperoxidase antineutrophil cytoplasmic antibody-positive group (n = 20); SS-A, anti-Sjögren’s syndrome A antibody-positive group (n = 20); Suspected, suspected COVID-19 group (n = 53); Positive, COVID-19-positive group (n = 26).

### Concordance rate with PCR testing

To investigate clinical usefulness, we compared the results of the serological antibody tests with those of PCR tests. Among the 26 PCR-positive COVID-19 cases, 8 cases (30.8%) had IgM-positive results and 19 cases (73.1%) had IgG-positive results. Among the 53 PCR-negative COVID-19-suspected cases, 52 cases (98.1%) had IgM levels below 10 AU/mL and all the cases (100%) had IgG levels below 10 AU/mL (Table 2).

**Table 2.** Concordance rate between the results of PCR testing and SARS-CoV-2 IgM or IgG serological testing.

|  |  | SARS-CoV-2 PCR test |  |
| --- | --- | --- | --- |
|  |  | positive cases (n = 26) | negative cases (n = 53) |
| SARS-CoV-2 IgM | >10 AU/mL | 8 (30.8%) | 1 |
|  | ≤10 AU/mL | 18 | 52 (98.1%) |
|  |  | SARS-CoV-2 PCR test |  |
|  |  | positive cases (n = | negative cases (n = |
|  |  | 26) | 53) |
| SARS-CoV-2 IgG | >10 AU/mL | 19 (73.1%) | 0 |
|  | ≤10 AU/mL | 7 | 53 (100%) |

We measured SARS-CoV-2 IgM and IgG in PCR-positive subjects (n = 26) and PCR-negative subjects (n = 53). An antibody titer above 10 AU/mL was regarded as positive, according to the manufacturer's cutoff.

### Suspected false-positive IgM results might be caused by reactivity to N protein

In this study, we observed 5 suspected false-positive IgM results. As described in the *Materials and Methods* section, the measurement system tests the reactivity to both the N protein and the S protein of SARS-CoV-2. We investigated the reactivity of the samples to N protein and S protein separately, and only reactivity to N protein was observed in the 5 suspected false-positive samples (Table 3).

**Table 3.**
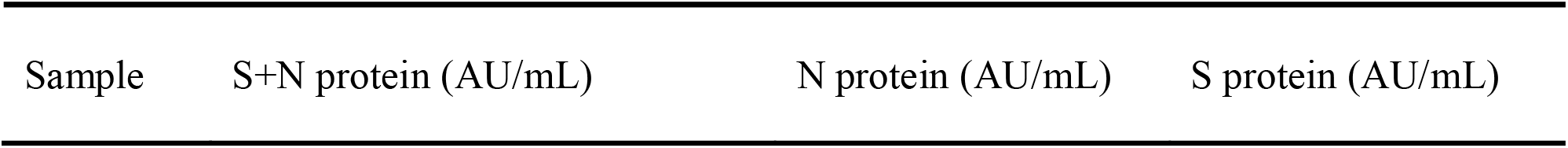
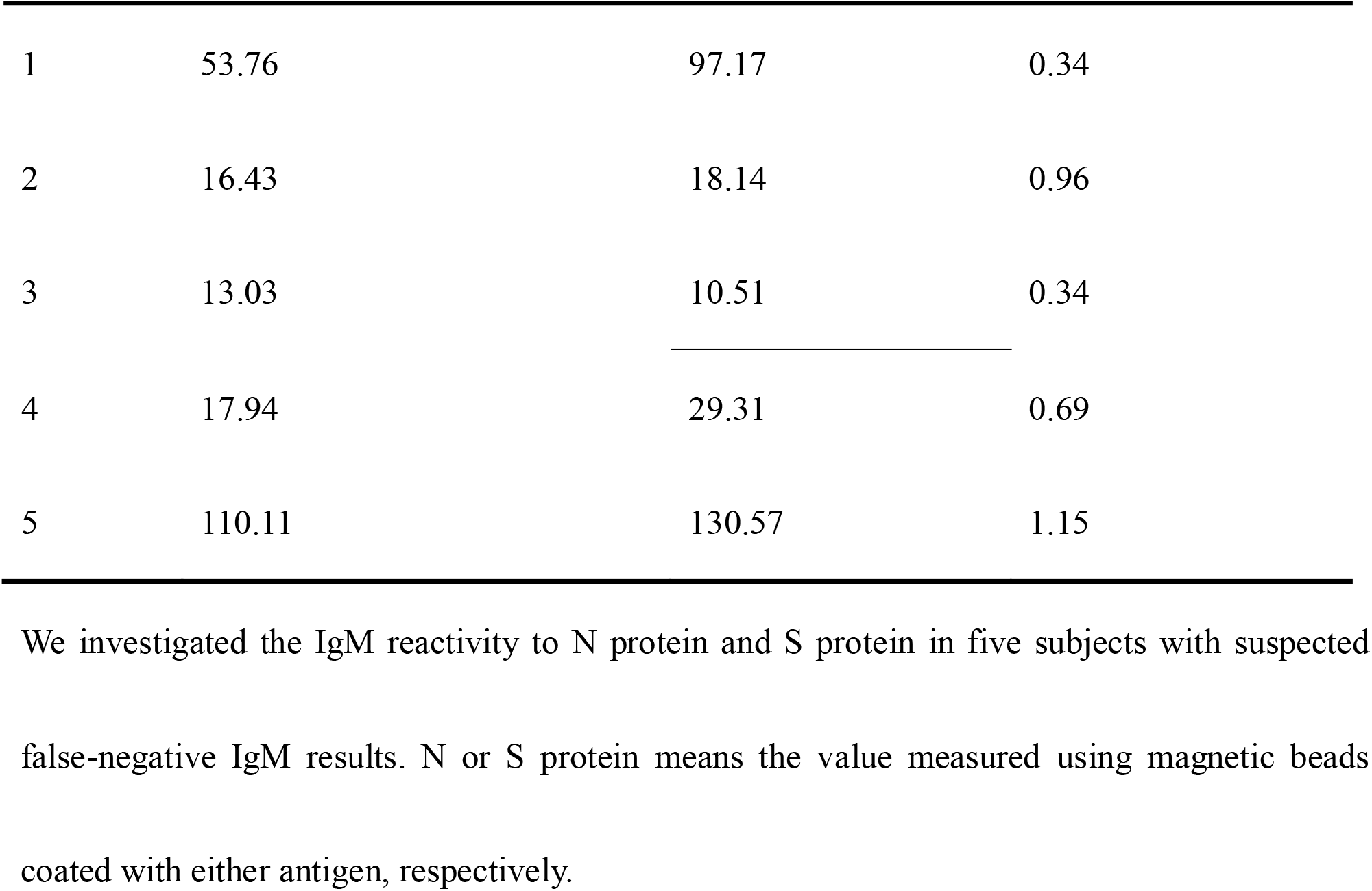
IgM reactivity to N protein and S protein in subjects with suspected false-negative IgM results.

**IgM results**
| Sample | S+N protein (AU/mL) | N protein (AU/mL) | S protein (AU/mL) |
| --- | --- | --- | --- |
| 1 | 53.76 | 97.17 | 0.34 |
| 2 | 16.43 | 18.14 | 0.96 |
| 3 | 13.03 | 10.51 | 0.34 |
| 4 | 17.94 | 29.31 | 0.69 |
| 5 | 110.11 | 130.57 | 1.15 |

We investigated the IgM reactivity to N protein and S protein in five subjects with suspected false-negative IgM results. N or S protein means the value measured using magnetic beads coated with either antigen, respectively.

## Discussion

In this study, we validated a method for quantifying SARS-CoV-2 IgM and IgG using the iFlash3000 automated CLIA analyzer. First, the within-day precisions of the SARS-CoV-2 IgM and IgG assays were obviously good in validations using three kinds of serum samples: a sample with a negative antibody titer, a sample with a titer close to the cutoff level, and an obviously positive sample (Table 1A). The between-day precisions for both assays also seemed good (Table 1B). These results suggest that relatively stable data were provided by this measurement system, while some anti-SARS-CoV-2 antibody detection kits authorized for emergency use by the Food and Drug Administration (FDA) in the USA have a CV of more than 5% for within-day precision and some manufacturers do not even publish precision data for their products. The linearity was good up to values of 112.7 AU/mL for SARS-CoV-2 IgM and 55.3 AU/mL for IgG, while the assay signal gradually reached a plateau at over 2,405 AU/mL for SARS-CoV-2 IgM and over 73 AU/mL for IgG (Fig. 2). Therefore, the sample should be diluted if the measured value is expected to be higher than the measurement range. To dilute samples, non-reactive serum should be used as a diluent, since the titers are calculated as values lower than the actual values when saline is used as a diluent, especially for IgM measurements (data not shown). Clinical decisions are rarely affected by this phenomenon, since the measurement range that was validated in the present study covers much higher values than the manufacturer’s cutoff.

Regarding the antibody tests, in addition to issues surrounding accuracy, the matter of false-positive cases has also been a concern. As shown in Fig. 3, the SARS-CoV-2 IgM/IgG levels were negative before hospitalization in three cases, and these antibody levels subsequently became positive after symptom onset in all cases, suggesting that this serological test can surely detect antibodies against SARS-CoV-2. In addition, we demonstrated that this measurement system could detect SARS-CoV-2 IgM/IgG without any false-positive results by evaluating 100 serum samples collected in 2017, when SARS-CoV-2 did not exist. These results confirm that this measurement system might be able to detect antibodies against SARS-CoV-2 alone, without cross-reactivity with other coronavirus strains that cause 15%-29% of all common colds.[20,21] Chemiluminescence immunoassays are known to be affected by autoantibodies, such as rheumatoid factor, relatively often.[22] In the present study, we investigated whether five kinds of autoantibodies might interfere with the measurement system and found that no false-positive results were observed for the IgG assays, while four false-positive cases were observed for the IgM assays. We also found one PCR-negative case with a SARS-CoV-2 IgM titer over the cutoff value. Actually, when we investigated the reactivity of N protein and S protein to the sera separately, we found that the false-positive cases were caused by reactivity to the N protein (Table 3). The reason for this reactivity remains unclear at present but might be due to the cross-reactivity of the measurement system or, since the structure of the N protein of SARS-CoV-2 is similar to that of other coronavirus strains, antibodies to a structure similar to the N protein of SARS-CoV-2 might actually exist.

Finally, we investigated the concordance rate between PCR and this measurement system. As shown in Table 2, although all the PCR-negative subjects had negative results except for one subject with a high IgM level, the PCR-positive subjects did not necessarily have positive results for IgM or IgG. This discrepancy might be due to the fact that we used the serum sample that had been collected on the day nearest to the day on which the PCR sample had been collected. In several cases, only a few days had passed since the onset of symptoms, and IgG and IgM are reportedly not detectable during the early phase of COVID-19.[23] Therefore, the time course for the appearance of IgG and IgM must be investigated for the application of antibody tests in clinical practice.

A characteristic of the present method is that both the N protein and the S protein are used as antigens. Reportedly, the sensitivity and specificity might depend on the types of antigens that are recognized by the antibodies, and the antibody to S protein is more sensitive than the antibody to N protein.[24–26] Since the measurement system in the present study uses both the S and N proteins, this system might provide a greater sensitivity and diagnostic ability than an antibody test using either the S protein or the N protein alone. In contrast, analyzing the reactivity to both proteins could increase the number of false-positive cases, as described above. Further studies on the clinical significances of antibodies to N protein and S protein might be necessary to conclude which is most appropriate: measuring the reactivity to both proteins or to each protein separately.

In conclusion, we have validated a measurement system for detecting IgM and IgG against SARS-CoV-2 using CLIA kits and have observed that this system had sufficient performance for its introduction into clinical laboratory testing. Moreover, the possibility of false-negative results, especially for IgG against SARS-CoV-2, was relatively low. In the future, this method might be helpful for clinical diagnosis, epidemiological investigations, and the development of vaccines.

## Data Availability

The datasets generated or analyzed in the current study are available upon reasonable request.

## Acknowledgements

We are thankful to the Murakami Foundation for the donation of the iFlash3000 to The University of Tokyo Hospital.

## Authorship Contributions

R.Y. participated in experiments and the data analysis and drafted the initial manuscript, Y.N., Y.M., T.S., Y.N., and N.Y. participated in the experiments. M.K. participated in the study design, data analysis, helped to draft the manuscript, and conceived the study. C.Q., F. X., F. H., Y. K. and J. O. developed the antibody measurement system. H.O., K.M., and Y.S. participated in the discussion and helped to draft the manuscript. T.K. and Y.Y. conceived the study, coordinated the study design, and helped to draft the manuscript. All the authors have read and approved the final manuscript.

## Data Availability Statement

The datasets generated or analyzed in the current study are available upon reasonable request.

### Abbreviations

AU: arbitrary unit
CLIA: chemiluminescence immunoassay
COVID-19: coronavirus disease 2019
CV: coefficient variation
dsDNA: double-strand DNA
FDA: the Food and Drug Administration
IgG: immunoglobulin G
IgM: immunoglobulin M
M2: mitochondrial M2
N protein: Nucleocapsid protein
P-ANCA: myeloperoxidase antineutrophil cytoplasmic antibody
RF: rheumatoid factor
RLU: relative light units
SARS-CoV-2: severe acute respiratory syndrome coronavirus 2
SD: standard division
S protein: Spike protein
SS-A: Sjögren’s syndrome A

